# Diagnosis of Blood Diseases and Disorders with Topological Deep Learning

**DOI:** 10.1101/2025.01.21.25320908

**Authors:** Philmore F. Koung, Saba Fatema, Nagehan Pakasticali, Hung S. Luu, Baris Coskunuzer

## Abstract

Blood diseases and disorders, including leukemia and infectious diseases of red blood cells, pose significant diagnostic challenges due to their complex presentations and reliance on time-consuming cytomorphological analysis to detect subtle morphologic features. While microscopic examination remains the gold standard in their diagnosis, its dependence on expert interpretation highlights the need for advanced methods to enhance the diagnostic workflow. Recently, deep learning (DL) methods have shown promise in medical imaging by automating and improving accuracy. However, these approaches often require large, annotated datasets, which are scarce for rare diseases, and they often face interpretability issues, limiting their integration into clinical practice.

In this study, we present a novel framework that integrates topological features with state-of-the-art DL techniques to enhance the analysis of cytomorphological images for diagnosing blood disorders. By combining global topological information with the localized patterns captured by DL models, our approach improves diagnostic accuracy while ensuring robustness, interpretability, and reproducibility. Experimental results demonstrate that the inclusion of topological features not only enhances model performance but also proves particularly effective in limited data settings. This methodology addresses critical limitations of existing techniques, advancing the classification and diagnosis of blood disorders while improving efficiency and reliability in medical imaging workflows.

## 1 Introduction

Blood diseases and disorders, including various types of blood cancers, represent a significant global health burden, often characterized by complex diagnostic processes and variable clinical presentations. Accurate diagnosis and classification of these entities rely on cytomorphology and histopathology, including detailed microscopic examination of blood smears and bone marrow samples. While this method remains the gold standard for the diagnosis, it is inherently time consuming and highly dependent on the expertise of the pathologists. With an increasing demand for accurate and timely diagnostic methods, the limitations of traditional approaches highlight the urgent need for advanced technologies to streamline and augment pathological workflows.

Recent breakthroughs in machine learning (ML) and deep learning (DL) have opened new avenues in medical imaging, providing opportunities to enhance diagnostic capabilities. DL models, with their ability to detect intricate patterns in images, have tremendous potential for automating and improving histopathological analysis. However, these models often face challenges such as the need for extensive labeled datasets, which are particularly difficult to curate in the context of rare diseases due to their low prevalence and limited availability of annotated data.

To overcome these issues, we propose the integration of topological data analysis (TDA) with DL frameworks for the analysis of histopathological images in rare blood diseases and blood cancers. TDA offers a unique perspective by capturing the topological structures and global features in images, which complement the localized patterns typically identified by DL models. By combining these global and local insights, the proposed methodology aims to create more robust, interpretable, reproducible and accurate diagnostic models, ultimately enhancing the effectiveness of the diagnosis and classification of blood diseases.

Our contributions can be summarized as follows:

- We introduce topological machine learning methods to cytomorphology, providing a novel approach to analyzing cellular structures in critical blood disorders.
- We show that topological features are highly effective in diagnosis of various blood disorders, including acute lymphoblastic leukemia (ALL), acute myeloid leukemia (AML), malaria, and babesiosis.
- We demonstrate that integrating topological feature vectors with DL models significantly enhances the performance, making these models more robust and reliable.
- In limited data settings where DL models typically struggle, topological vectors alone achieve strong results and substantially boost the performance of DL models.
- We highlight the interpretability of topological features, addressing a crucial need for leveraging black-box ML models in this domain.

## 2 Related Work

### 2.1 Machine Learning Methods for Rare Blood Disease Diagnosis

Machine learning (ML) and deep learning (DL) have emerged as transformative tools in detecting and diagnosing blood disorders through histopathological and cytomorphological image analysis. These techniques enhance the ability to identify subtle pathological patterns, improve diagnostic accuracy, and assist clinicians in decision-making. Notable works include the DL-based models in [27] and [35], which demonstrated expert-level performance in classifying acute myeloid leukemia (AML) and acute lymphoblastic leukemia (ALL) using high-resolution cytomorphological and blood smear images. To address the scarcity of annotated data for rare diseases, transfer learning has been effectively employed. For instance, [24] integrates pre-trained CNN architectures like ResNet and DenseNet to classify subtypes of myelodysplastic syndromes (MDS) and other rare hematologic conditions, leveraging general medical imaging datasets to improve model performance.

Recent studies have also explored multimodal frameworks and feature selection techniques to enhance model interpretability and diagnostic accuracy. Multimodal approaches, such as [22] and [44], integrate cytomorphological image data with genomic and clinical information to improve the diagnosis and risk stratification of complex blood disorders. Feature selection techniques, like those used in [25], refine extracted features from cytological images, optimizing classification performance for various hematological malignancies. Comprehensive reviews, such as [39] and [16], highlight the progress and challenges in applying ML and DL methods to rare blood diseases, emphasizing their potential to bridge clinical gaps. These advancements demonstrate the promise of ML and DL in transforming diagnostic workflows and improving patient outcomes in hematological oncology.

### 2.2 TDA in Medical Image Analysis

Cubical persistence has proven to be a powerful tool for extracting topological features in medical image analysis, with applications in tumor segmentation [33], brain connectivity studies [6,36], glioblastoma outcome prediction [10], thoracic disease diagnosis [2] and genomic data exploration [34]. Resources like the TDA Applications Library [14] and DONUT [15], alongside recent surveys [38], further highlight its adaptability. Advances in scalable computational frameworks [4,12] have enabled the application of persistent homology (PH) to large-scale biomedical datasets.

The integration of PH with machine learning has unlocked new possibilities in diagnostic workflows. PH has been incorporated to enhance image segmentation frameworks [17,37,41] and to predict treatment responses in breast cancer [42]. Recent advancements in topological deep learning underscore its ability to augment CNN-based models for segmentation, classification, and precision medicine applications [32,45]. For instance, PH has been utilized for leukemia classification and the diagnosis of rare blood disorders, offering novel insights into cytomorphological imaging [40,43].

## 3 Methodology

Our methodology involves two key steps. First, we extract topological feature vectors from the images. Next, we leverage these vectors to diagnose blood disorders by effectively integrating them with recent DL models.

### 3.1 Topological Vectors for Images

Persistent Homology (PH) is a foundational tool in topological data analysis (TDA) that captures the underlying shape and structure of complex datasets. By systematically tracking the emergence and disappearance of topological features at different resolutions, PH provides a robust framework for understanding patterns in diverse data types, including point clouds, networks, and images [7]. Here, we focus on the application of *cubical persistence*, a variant of PH tailored specifically for analyzing image data.

While we aim to present the construction and application of PH in an accessible manner, detailed theoretical discussions can be found in [11,9]. Here, we outline the process of using cubical persistence for feature extraction from images through three main steps: (1) constructing filtrations, (2) generating persistence diagrams, and (3) vectorizing the persistence information for use in ML models.

#### Step 1: Constructing Filtrations

Filtration is the process of building a sequence of nested topological spaces that represent the progressive inclusion of features in the dataset. For image analysis, this involves constructing *cubical complexes*—a specific type of topological structure derived from binary images.

From a grayscale or color image 𝒳 (dimensions *r* × *s*), we extract pixel intensities *γ*_*ij*_ for each pixel *Δ*_*ij*_ ⊂𝒳. A *sublevel filtration* is constructed by sequentially activating (coloring black) pixels as their grayscale values reach predefined thresholds *t*_1_ *< t*_2_*<*···*< t*_*N*_ = 255. This creates a sequence of binary images 𝒳_1_ ⊂ 𝒳_2_ ⊂ · · · ⊂ 𝒳_*N*_, where 𝒳_*n*_ = {*Δ*_*ij*_ ⊂𝒳 |*γ*_*ij*_ ≤*t*_*n*_}. Alternatively, a *superlevel filtration* can be constructed by deactivating pixels in descending order of intensity. This results in another sequence, 𝒴_1_ ⊂ 𝒴_2_ ⊂ · · · ⊂ 𝒴_*M*_, where 𝒴_*n*_ = {*Δ*_*ij*_ ⊂ 𝒳 | *γ*_*ij*_ ≥*s*_*n*_}, with *s*_1_ *> s*_2_ *>* · · ·*> s*_*M*_ = 0. Both filtrations provide complementary views of the image’s topology, enabling comprehensive feature extraction.

#### Step 2: Persistence Diagrams

PH captures the evolution of topological features (e.g., connected components, loops, cavities) across the filtration. This information is recorded in a *persistence diagram (PD)*, which maps each feature *σ* to its birth and death thresholds (*b*_σ_, *d*_σ_). For a sublevel filtration {𝒳_*n*_}, *Birth time b*_σ_ marks the threshold *t*_*m*_ where a feature *σ* first appears, and *Death time d*_σ_ indicates the threshold *t*_*n*_ where *σ* disappears. The persistence diagram PD_*k*_(𝒳) for dimension *k* contains all such pairs for *k*-dimensional features (e.g., PD_0_ for connected components, PD_1_ for loops). Features with long lifespans (*d*_σ_−*b*_σ_) are typically more significant, as they represent prominent topological structures. For instance, in the filtration example in Figure 2, PD_0_(𝒳) captures connected components, while PD_1_(𝒳) records loops (holes).

**Fig 1:**
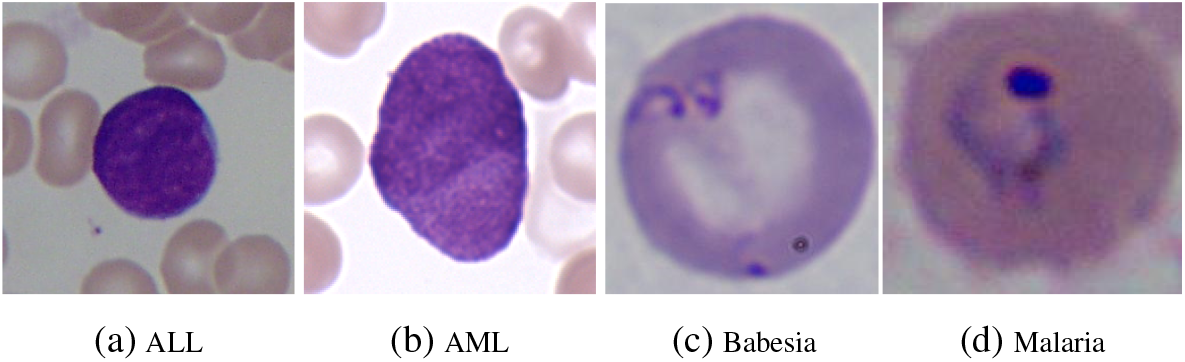
Sample single cell images for ALL, AML, Malaria and Babesia.

**Fig 2:**
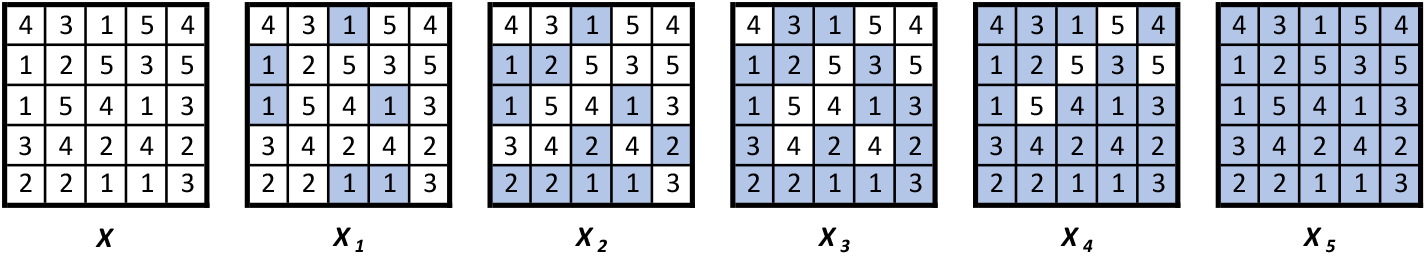
For the 5 × 5 image 𝒳 with the given pixel values, **the sublevel filtration** is the sequence of binary images 𝒳_1_ ⊂ 𝒳_2_ ⊂ 𝒳_3_ ⊂ 𝒳_4_ ⊂ 𝒳_5_.

**Fig 3:**
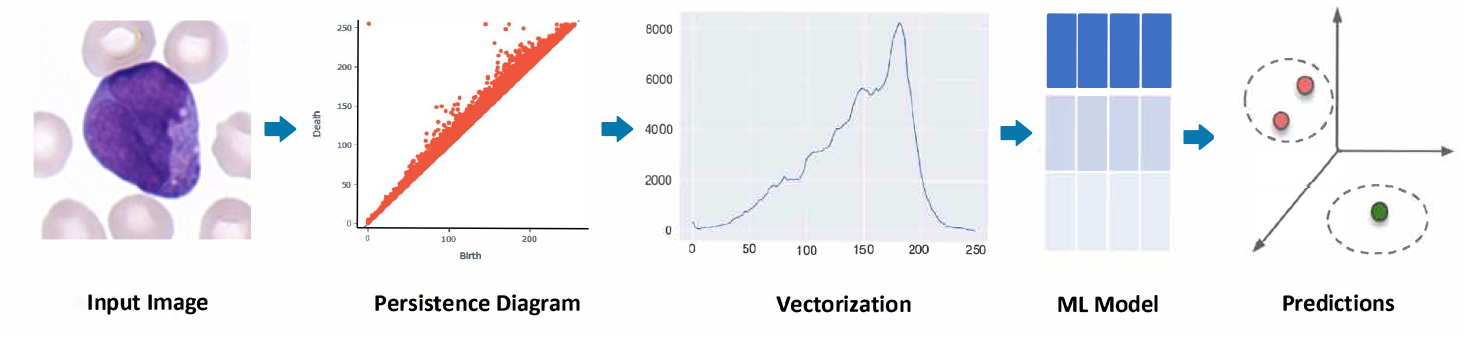
Topo-Vita Model. In our basic topological model, we first extract topological feature vectors for each image through cubical persistence and feed them into standard ML models.

#### Step 3: Vectorization for Machine Learning

Persistence diagrams, while informative, are collections of 2D points that are challenging to use directly in ML models. Thus, vectorization techniques are employed to convert persistence diagrams into a fixed-dimensional representation. One widely used approach is the *Betti function*, which tracks the number of alive features at each threshold. For an image 𝒳, the Betti function *β*_*k*_(*t*) at threshold *t* is defined as the count of *k*-dimensional features in 𝒳_*n*_. This function can be represented as a vector 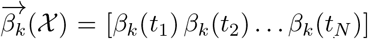. For example, in Figure 2, we get 5-dimensional vectors, 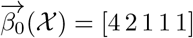 and 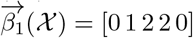.

Other advanced vectorization methods, such as persistence landscapes [5], persistence images [1], and kernel-based approaches [3], offer alternatives with varying computational and interpretive benefits [3]. However, we emphasize Betti functions in this study due to their efficiency and interpretability.

### 3.2 ML Models

In this study, we propose a novel approach to diagnosing blood disorders by integrating topological vectors with DL models. For each image in our dataset, we generate a 400-dimensional feature vector that captures its topological characteristics (Section 3.1). These vectors can be utilized independently or in conjunction with the medical images to train ML models. To evaluate the utility of these features, we design two distinct types of models, each exploring different aspects of their integration and performance.

#### Topo-Vita: Evaluating Standalone Topological Features

This is our basic model to initially test the effectiveness of our topological vectors for classifying blood disorders. We simply incorporate two standard ML models with our topological vectors.

##### Multilayer Perceptron

A flexible feedforward neural network that has demonstrated robustness in handling diverse feature sets. By using MLP, we assess how well topological vectors alone can capture the diagnostic information embedded in the images.

##### TabTransformer

A transformer-based model designed specifically for tabular data [21]. Given that our topological features can also be interpreted as *sequential data*, the Tab-Transformer provides an effective mechanism to explore their potential in a more structured and context-aware manner.

#### Topo-VitaX: Enhancing DL Models with Topological Features

Our advanced model evaluates the impact of integrating topological vectors into pre-trained DL architectures. The rationale behind this approach is that topological features capture global patterns within an image, offering embeddings that remain robust regardless of data quantity. In contrast, convolutional layers emphasize localized patterns, with their performance heavily reliant on the volume of available data. To explore this synergy, we employed two distinct types of DL architectures.

##### Pre-trained CNNs

We purposefully choose a simple architecture (Figure 4) to isolate and highlight the contribution of topological features in improving DL models for classifying rare blood disorders. The fusion involves concatenating the topological vectors with the CNN’s feature embeddings before the final classification layers.

**Fig 4:**
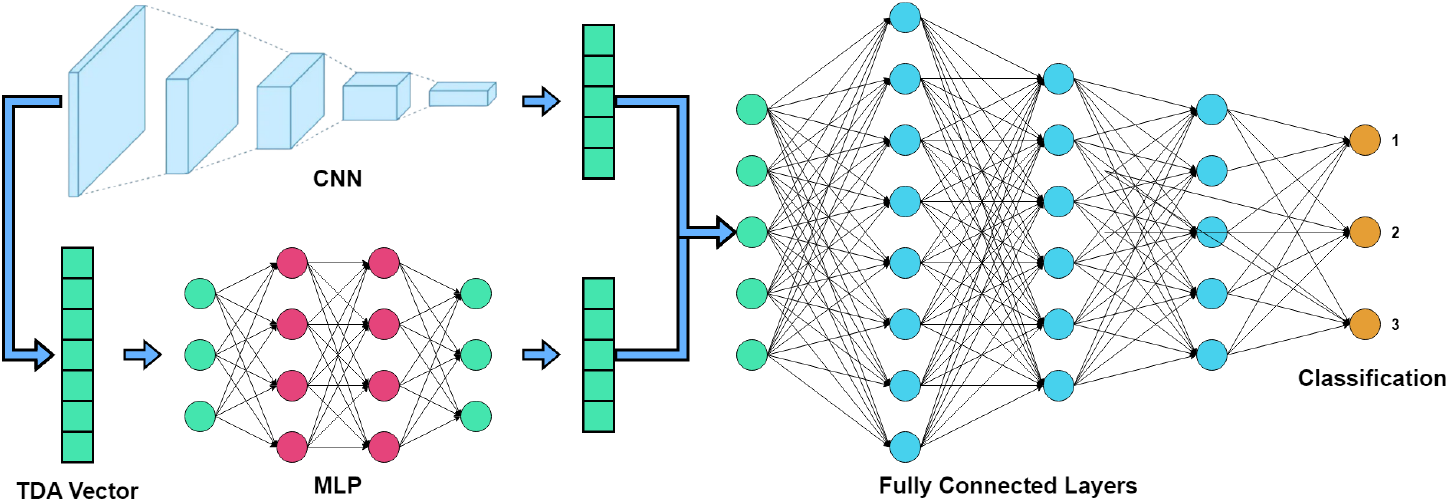
Topo-VitaX Model. In our augmented CNN, we integrate our topological features of images obtained in the Topo-Vita model with convolutional vectors generated by a CNN backbone, followed by processing through a fully connected layer.

##### Vision Transformers (ViTs)

In our approach, topological vectors are directly fed into the attention mechanism of ViTs as an additional input, enabling the model to leverage these global structural features during the attention computation. This integration allows the topological features to influence the contextual representation learning within the ViT, enhancing its ability to classify complex patterns in blood smear images.

We give the details of our backbone DL models and hyperparameters in Section 4.1.

## 4 Experiments

### 4.1 Experimental Setup

#### Datasets

For our experiments, we aimed to classify single-cell images of blood disorders, including leukemia and infectious diseases of red blood cells, distinguishing them from normal cells. Specifically, we focused on acute lymphoblastic leukemia (ALL), acute myeloid leukemia (AML), Babesiosis, and Malaria (See Figure 1 for image samples). We used the following publicly available datasets.

For ALL, we utilized the ALL-IDB2 dataset from the Acute Lymphoblastic Leukemia Image Database (ALL-IDB) [23]. For AML, we employed the AML-Cytomorphology MLL Helmholtz dataset from the Cancer Imaging Archive [19], which includes four subclasses: (i) Acute Promyelocytic Leukemia (APL) with PML::RARA fusion, (ii) AML with NPM1 mutation, (iii) AML with CBFB::MYH11 fusion (excluding NPM1 mutation), and (iv) AML with RUNX1::RUNX1T1 fusion, along with a control group. Each class comprises 1000 images.

We conducted experiments using two setups: binary classification (normal vs. abnormal) and 5-way multi-class classification. For Malaria, we relied on the Malaria dataset from the National Library of Medicine [30]. To differentiate between Malaria, Babesiosis, and noninfected cells, we used the Microscopic Images of Parasite Species dataset from the National Institute of Allergy and Infectious Diseases (NIAID) [29]. For all datasets, we applied a 70:10:20 random split for training, validation, and testing, respectively.

#### Topological Vectors

For all of our datasets, we initially generate the topological feature vectors. First, we apply sublevel filtration using 50 evenly spaced thresholds from 0 to 255 for each color channel (red, blue, green, and grayscale). Following this step, we then apply Betti vectorization to the resulting persistence diagrams and concatenate the resulting vectors for each color channel, creating a 400-dimensional feature vector (or Betti vector) for each image in each dataset. Figure 7 illustrates how our topological vectors distinguish different class in blood disorder datasets.

#### ML Model Hyperparameters

To test the standalone effectiveness of Betti vectors, we trained an MLP on 300 epochs with early stopping monitoring validation loss and patience set to 5. Our model consists of 3 hidden layers, the first hidden layers containing 256 units with the last 2 layers containing 128 units. We used ReLU activation functions, cross-entropy loss, and the Adam optimizer with learning rate set to 0.0001 initially, then multiplied by 0.1 after the 50th epoch, and multiplied again by 0.1 after the 75th epoch. Our TabTransformer model was also trained on 300 epochs with dimension embedding of 128, 4 heads, and 5 layers. Additionally, we used the same learning rate scheduler and early stopping setup.

#### DL Model Hyperparameters

We used a batch size of 128 for all datasets with 128×128 images for the AML, Malaria, and Babesia datasets and 224×224 images for the ALL dataset. Again, we used cross-entropy loss with the Adam optimizer with a learning rate of 0.0001 multiplied by 0.1 after the 50th epoch and multiplied by 0.1 again after the 75th epoch. Key performance metrics are incorporated such as accuracy and AUC. For our auxiliary Betti vector MLP we used 3 hidden layers, the first with 256 units and 128 units for the last 2 layers. Our classifier MLP consists of 2 hidden layers with 256 units each. We selected feature dimensions of 128 for both the CNN and Betti MLP architectures, based on comparative evaluations of different feature size ratios in our earlier experiments. All MLPs and CNNs used ReLU activation functions. Similar to our ML model, we trained our DL models on 300 epochs and used early stopping monitoring validation loss with patience set to 5. Our code is available at ^3^.

#### Baseline Models

To evaluate the effect topological features have when combined with DL models, we first choose a base model from a range of CNN models. We used ResNet50 [18], DenseNet121 [20], and Xception [8] as baselines. We also include a vision transformer model, Swin Transformer V2-Tiny [26]. These models were chosen due to their robust feature extraction capabilities and performance on image classification tasks. The details of our architectures are given in Section 3.2.

#### Computational Complexity & Runtime

The computation of topological vectors can be resource-intensive, particularly when applied to high-dimensional datasets [31]. For 2D images, however, the process is relatively efficient, with a time complexity of approximately 𝒪 (|𝒫|^2.3^), where |𝒫| represents the total number of pixels [28]. This implies that the computational demand increases nearly quadratically with the size of the image. By comparison, subsequent tasks such as vectorization and machine learning are significantly less computationally demanding.

We conducted all experiments on a high-performance computing system containing one Nvidia L40S. The L40S contains 8 vCPU, 62 GB RAM, and 48 GB VRAM. Generating both Betti-0 and Betti-1 vectors from the Malaria dataset, which contains 27,558 images of size 128 × 128, took 2 hours 32 minutes and 55 seconds. The remaining ML tasks were significantly faster. The most complex model T-SwinV2 for Malaria was completed in 17 minutes and 32 seconds. For the remaining datasets, the generation of topological vectors and the runtime of the ML models were significantly faster.

#### Performance Metrics

We used widely recognized performance metrics in the biomedical domain, including Accuracy, ROC-AUC, Precision, and Recall. Given the challenges posed by unbalanced datasets, we primarily focus on reporting ROC-AUC results across most settings. However, detailed results for all metrics are given in Table 4.

### 4.2 Results

This study had two primary objectives. The first was to demonstrate the effectiveness of topological vectors in detecting blood disorders from blood smear images. To achieve this, we developed a basic ML model that combines topological vectors with traditional ML techniques, allowing us to evaluate the standalone performance of topological vectors. The second objective was to assess the impact of integrating topological vectors with state-of-the-art deep learning models to enhance their performance.

#### Standalone Performance of Topological Vectors

The first column of Table 2 and the first two columns of Table 4 present the standalone performance of our topological vectors (Topo-Vita model). It is important to note that this simple model does not involve any pre-training and relies solely on topological features. Across all datasets, the performance is notable, with topological vectors outperforming most DL models and achieving competitive, head-to-head results with the rest.

**Table 1:**
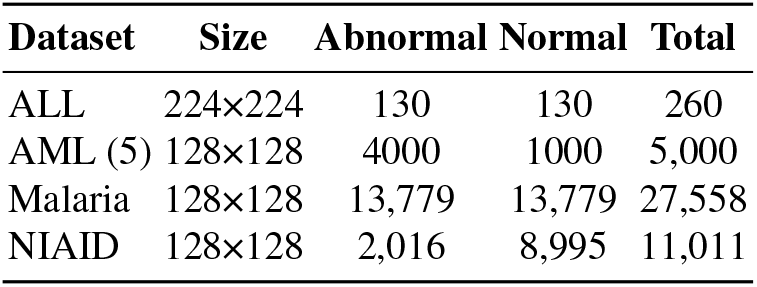
Dataset Statistics.

| Dataset | Size | Abnormal | Normal | Total |
| --- | --- | --- | --- | --- |
| ALL | 224×224 | 130 | 130 | 260 |
| AML (5) | 128×128 | 4000 | 1000 | 5,000 |
| Malaria | 128×128 | 13,779 | 13,779 | 27,558 |
| NIAID | 128×128 | 2,016 | 8,995 | 11,011 |

**Table 2:** ROC-AUC Results. Each dataset includes a control group as one class. The *Topo* column highlights the standalone performance of topological vectors, while the *T-Net models* represent DL models enhanced with these vectors. For each dataset, the highest improvement for T-Net models is marked blue, and the best-performing model is marked **bold**. The further performance metrics are given in Table 4.

| Disease | Class | Topo | ResNet | T-ResNet | DenseNet | T-DenseNet | Xception | T-Xception | Swin | T-Swin |
| --- | --- | --- | --- | --- | --- | --- | --- | --- | --- | --- |
| ALL | 2 | **1.0000** | 0.9568 | 0.9792 | 0.9799 | 0.9412 | **0.8683** | **0.9286** | 0.9231 | 0.9673 |
| AML | 2 | 0.9389 | 0.9245 | 0.7897 | **0.7956** | **0.8756** | 0.8646 | 0.9373 | 0.9040 | **0.9430** |
| AML | 5 | 0.8281 | 0.7894 | 0.7920 | 0.7522 | 0.7911 | **0.6950** | **0.8215** | **0.8718** | 0.8324 |
| Malaria | 2 | 0.9334 | **0.7213** | **0.9292** | 0.9481 | 0.9419 | 0.9345 | 0.9360 | 0.9688 | **0.9932** |
| Babesiosis | 2 | 0.9964 | 0.9984 | 0.9994 | 0.9976 | 0.9999 | **0.9832** | **0.9998** | 0.9968 | **1.0000** |
| Babes/Malaria | 3 | 0.9982 | 0.9992 | **0.9993** | 0.9986 | 0.9989 | **0.9946** | **0.9984** | 0.9973 | 0.9971 |

Additionally, the distinguishing capability of topological vectors is visualized in t-SNE projections (Figure 5). In Figure 5a, the topological vectors clearly separate the ALL and normal classes, achieving a perfect AUC score of 1.0. Furthermore, ALL images form a distinct cluster in the lower left corner, while normal classes are more dispersed. This pattern suggests the potential existence of multiple ALL subtypes, which could be explored in future studies. In Figure 5b, the separation between classes is less pronounced compared to the ALL dataset. However, this visualization is a 2D projection of a 400-dimensional latent space, and the very high AUC score highlights the strong discriminative power of topological vectors in the original latent space. Finally, in Figure 5c, we observe clear and well-defined clusters for normal, Babesia, and malaria classes, reflecting the robust performance of topological vectors with an AUC score of 0.99. This visualization underscores their effectiveness in distinguishing these categories.

**Fig 5:**
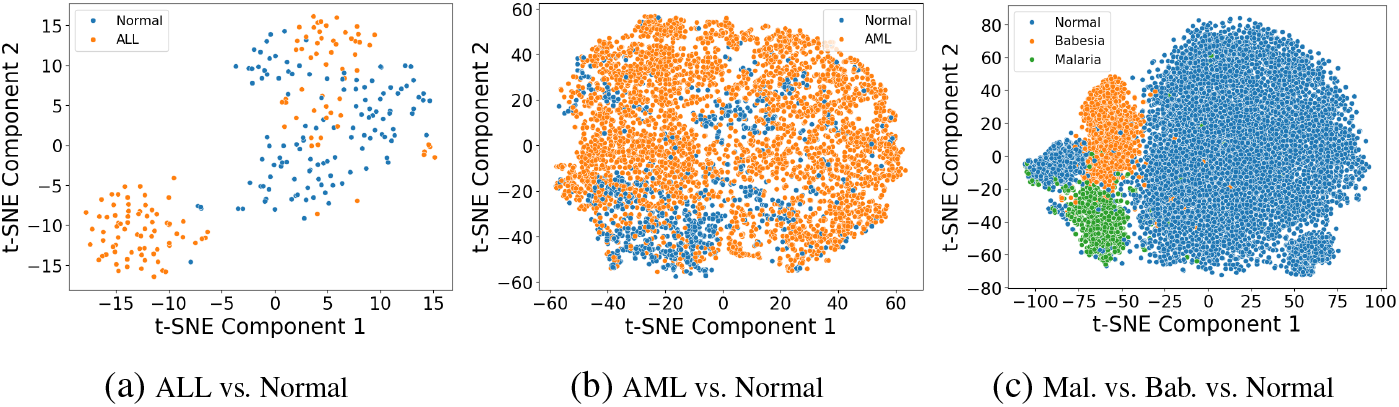
t-SNE Figures. For each image, we extract a 400-dimensional topological feature vector as described in Section 3. The figures above illustrate 2D t-SNE projections of these topological embeddings for each dataset, where each point represents an image.

**Fig 6:**
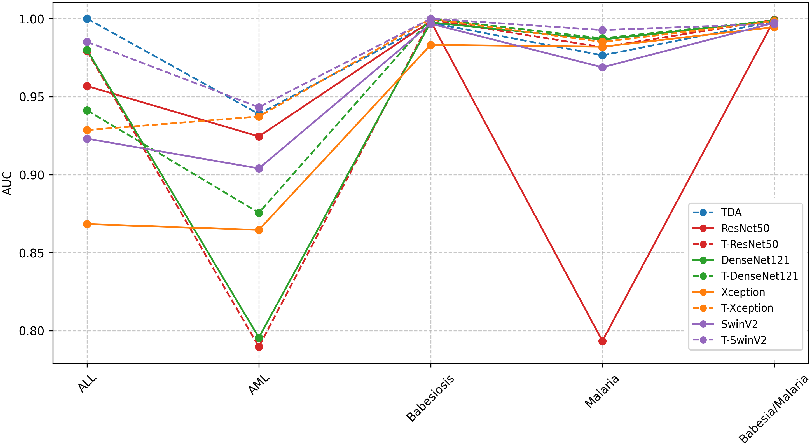
ROC-AUC results. The performance of DL models for each dataset is illustrated with solid lines, while topologically enhanced models are represented with dotted lines in matching colors. Detailed performance metrics are provided in Table 4.

**Fig 7:**
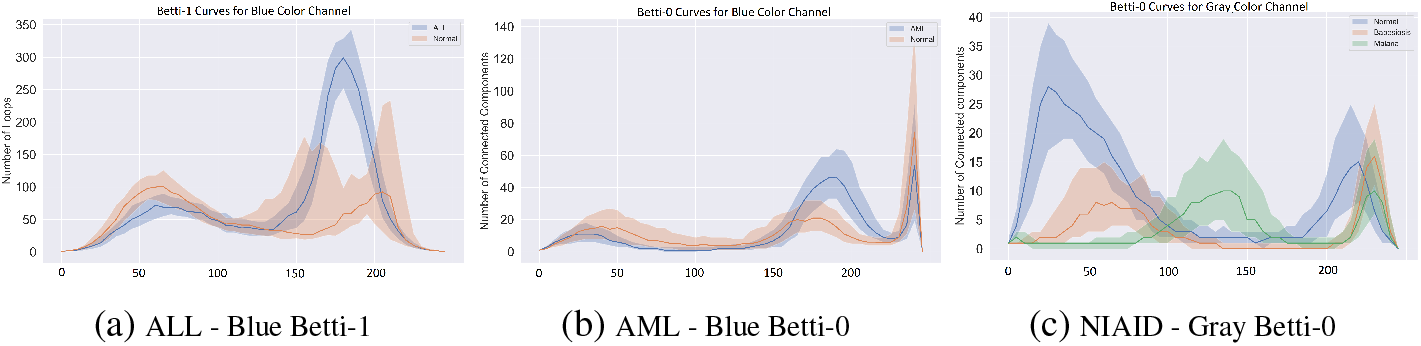
Betti Curves. 40% confidence bands around the median curves are shown for each class. The x-axis represents color values, while the y-axis indicates the count of components (Betti-0) and the count of holes (Betti-1) in the corresponding binary image *𝒳*_*t*_.

#### Improvements of DL Models with Topological Features

Our second objective was to evaluate the impact of incorporating topological vectors into popular deep learning (DL) models. Notably, all baseline models were pre-trained, meaning any observed improvement is particularly significant. As shown in Tables 2 and 4, the inclusion of topological vectors consistently enhances the performance of these pre-trained models, achieving up to a 20% improvement in AUC. These findings highlight the substantial potential for performance enhancement by integrating topological features with DL models. Even with a straightforward method of injecting topological features into CNN architectures, the results demonstrate meaningful gains. With domain-specific adaptations, this synergy between topological and DL features presents promising opportunities for further advancements in performance.

#### Limited Data Performance

One of the primary challenges deep learning (DL) models face is their reliance on large quantities of annotated data. In many medical domains, particularly in the context of rare diseases, labeled datasets are often scarce. While DL methods typically learn vector representations through backpropagation, topological methods generate direct embeddings by capturing the inherent topological patterns in the data. This approach makes topological embeddings less dependent on labeled data for effective performance. In this study, we assessed the utility of topological vectors in classifying blood disorders.

Table 3 presents the results of our experiments. For consistency, we held the 20% test set constant across all datasets and evaluated model performance using training subsets containing 10, 20, and 50 samples per class. To provide a fair comparison of learning performance, we utilized untrained, “vanilla” deep learning models rather than pre-trained ones. Our results demonstrate that in limited data settings, topological models consistently outperform DL models across all datasets. Furthermore, integrating topological vectors with DL models significantly enhances their performance, showcasing the complementary strengths of these approaches.

**Table 3:**
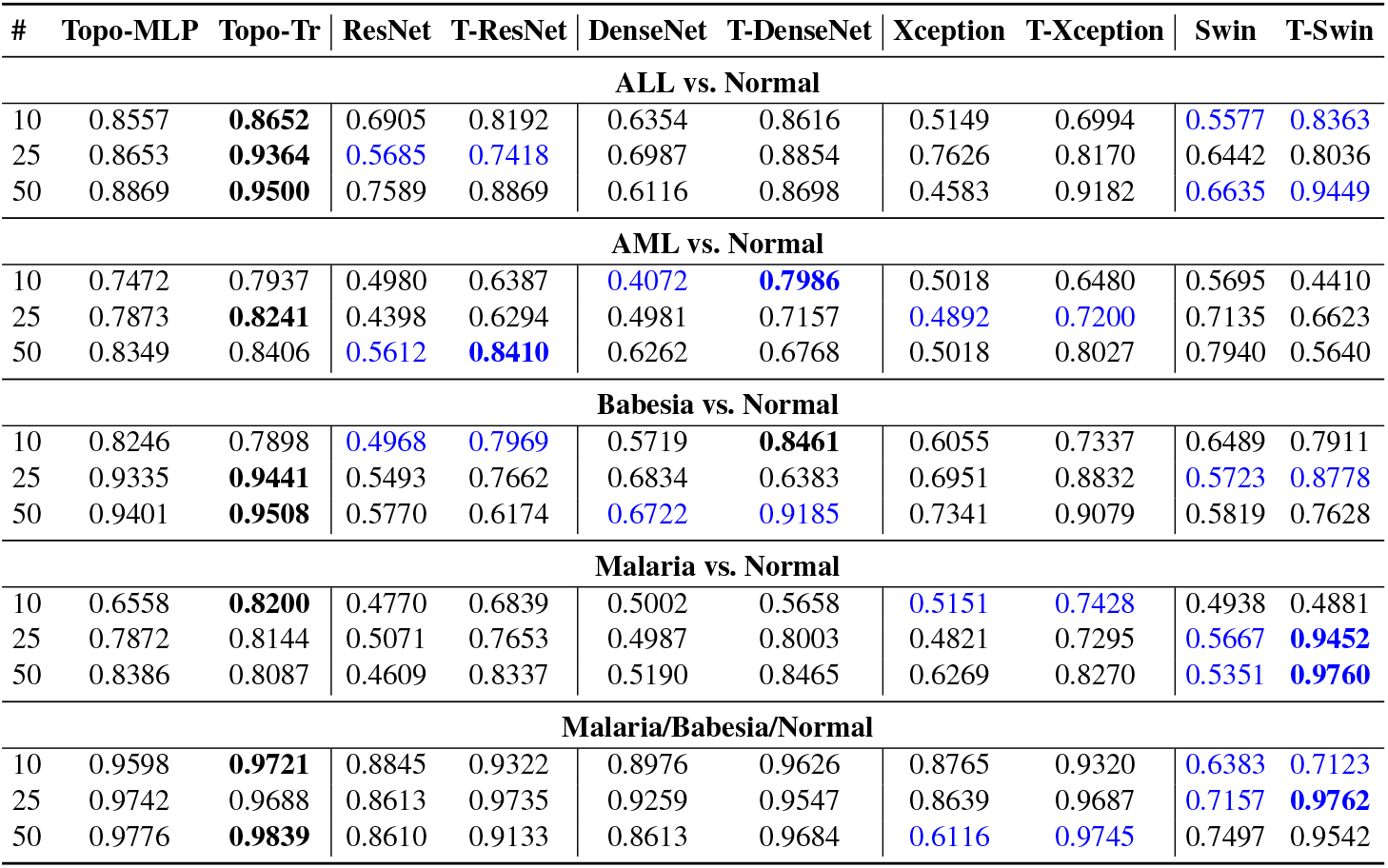
Limited Data Settings. AUC results of our models using 10, 25, and 50 training samples per class, with the original test set unchanged. For each row, the highest improvement for T-Net models is marked blue, and the best-performing model is marked **bold**.

**Table 4:** All Performance Metrics. Below we present all performance metrics for our models. The first two *Topo* columns show the standalone performance of topological vectors, while *T-Net models* represent DL models enhanced with these topological vectors.

| Metric | Topo-MLP | Topo-Tr | ResNet | T-ResNet | DenseNet | T-DenseNet | Xception | T-Xception | Swin | T-Swin |
| --- | --- | --- | --- | --- | --- | --- | --- | --- | --- | --- |
| **ALL vs. Normal** |  |  |  |  |  |  |  |  |  |  |
| Accuracy | 0.9038 | 0.9615 | 0.9038 | 0.9423 | 0.9038 | 0.9038 | 0.8077 | 0.8846 | 0.9231 | 0.9808 |
| AUC | 0.9397 | 1.0000 | 0.9568 | 0.9792 | 0.9799 | 0.9412 | 0.8683 | 0.9286 | 0.9231 | 0.9673 |
| Precision | 0.9259 | 1.0000 | 0.9259 | 0.9630 | 1.0000 | 0.9259 | 0.8750 | 0.8667 | 0.9231 | 1.0000 |
| Recall | 0.8929 | 0.9333 | 0.8929 | 0.9286 | 0.8214 | 0.8929 | 0.7500 | 0.9286 | 0.9231 | 0.9643 |
| **AML vs. Normal** |  |  |  |  |  |  |  |  |  |  |
| Accuracy | 0.8890 | 0.9000 | 0.8760 | 0.8080 | 0.8160 | 0.8720 | 0.8380 | 0.8840 | 0.9040 | 0.9160 |
| AUC | 0.9389 | 0.9138 | 0.9245 | 0.7897 | 0.7956 | 0.8756 | 0.8646 | 0.9373 | 0.9040 | 0.9430 |
| Precision | 0.9285 | 0.9205 | 0.9295 | 0.8290 | 0.8170 | 0.8962 | 0.8844 | 0.9185 | 0.9032 | 0.9383 |
| Recall | 0.9320 | 0.9627 | 0.9131 | 0.9575 | 0.9899 | 0.9500 | 0.9156 | 0.9370 | 0.9050 | 0.9572 |
| **AML (5) - 4 subtype & Normal** |  |  |  |  |  |  |  |  |  |  |
| Accuracy | 0.5130 | 0.5400 | 0.4990 | 0.4760 | 0.4240 | 0.4740 | 0.3670 | 0.5210 | 0.6190 | 0.5440 |
| AUC | 0.8235 | 0.8281 | 0.7894 | 0.7920 | 0.7522 | 0.7911 | 0.6950 | 0.8215 | 0.8718 | 0.8324 |
| Precision | 0.6980 | 0.5426 | 0.6347 | 0.6851 | 0.6396 | 0.6910 | 0.5962 | 0.6809 | 0.6505 | 0.5508 |
| Recall | 0.3120 | 0.5359 | 0.2850 | 0.2480 | 0.1970 | 0.2460 | 0.1240 | 0.3200 | 0.6190 | 0.5409 |
| **Babesia vs. Normal** |  |  |  |  |  |  |  |  |  |  |
| Accuracy | 0.9830 | 0.9809 | 0.9851 | 0.9915 | 0.9809 | 0.9894 | 0.9426 | 0.9809 | 0.9968 | 1.0000 |
| AUC | 0.9974 | 0.9964 | 0.9984 | 0.9994 | 0.9976 | 0.9999 | 0.9832 | 0.9998 | 0.9968 | 1.0000 |
| Precision | 0.9696 | 0.9793 | 0.9822 | 0.9911 | 0.9778 | 0.9612 | 0.9227 | 0.9614 | 0.9979 | 1.0000 |
| Recall | 0.9955 | 0.9833 | 0.9866 | 0.9911 | 0.9821 | 0.9955 | 0.9598 | 1.0000 | 0.9957 | 1.0000 |
| **Malaria vs. Normal** |  |  |  |  |  |  |  |  |  |  |
| Accuracy | 0.9334 | 0.9263 | 0.7213 | 0.9292 | 0.9481 | 0.9419 | 0.9345 | 0.9360 | 0.9690 | 0.9675 |
| AUC | 0.9765 | 0.9709 | 0.7934 | 0.9817 | 0.9866 | 0.9874 | 0.9820 | 0.9851 | 0.9688 | 0.9932 |
| Precision | 0.9499 | 0.9456 | 0.7067 | 0.9641 | 0.9604 | 0.9741 | 0.9500 | 0.9679 | 0.9692 | 0.9727 |
| Recall | 0.9159 | 0.9062 | 0.7623 | 0.8925 | 0.9354 | 0.9087 | 0.9181 | 0.9026 | 0.9690 | 0.9625 |
| **Malaria/Babesia/Normal** |  |  |  |  |  |  |  |  |  |  |
| Accuracy | 0.9832 | 0.9732 | 0.9918 | 0.9950 | 0.9868 | 0.9891 | 0.9619 | 0.9891 | 0.9976 | 0.9977 |
| AUC | 0.9982 | 0.9945 | 0.9992 | 0.9993 | 0.9986 | 0.9989 | 0.9946 | 0.9984 | 0.9973 | 0.9971 |
| Precision | 0.9841 | 0.9631 | 0.9927 | 0.9950 | 0.9868 | 0.9891 | 0.9627 | 0.9891 | 0.9964 | 0.9974 |
| Recall | 0.9828 | 0.9115 | 0.9918 | 0.9946 | 0.9868 | 0.9891 | 0.9610 | 0.9891 | 0.9964 | 0.9925 |

### 4.3 Interpretability of Topological Features

In this section, we delve into the interpretability of topological features, emphasizing Betti vectors as a simple yet powerful method for vectorizing persistence diagrams, prized for their high interpretability. Figure 7 illustrates the Betti curves for our datasets, augmented with nonparametric confidence bands [13]. These visualizations reveal significant differences in topological patterns across classes, underscoring the discriminative power of these features.

In the [0, 255] color scale, 0 represents black, and 255 corresponds to white. Thus, darker colors are activated earlier, while lighter regions are activated later. For a given threshold *t*, the binary image 𝒳_*t*_ includes all pixels *Δ*_*ij*_ with color values *γ*_*ij*_ ≤ *t*, meaning these pixels are activated (colored black).

In Figure 7a, we observe a distinct difference between the normal and ALL classes around the color value of 180. This is reflected in the *β*_1_(180) values, which represent the number of holes in the binary image 𝒳 𝒳_180_ for the blue color channel (Section 3.1). For the normal class, *𝒳* _180_ typically contains approximately 50 holes, whereas the ALL class exhibits around 300 holes. Figure 8 provides sample *𝒳*_180_ binary images for both classes, visually highlighting the significantly greater number of holes in the ALL image compared to the normal image.

**Fig 8:**
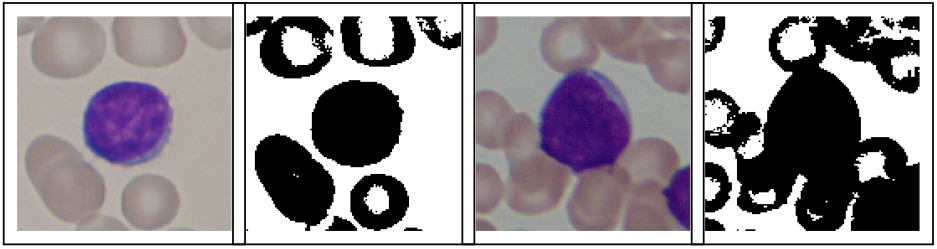
Sample single cell images and their binary 𝒳_180_ images for Normal (left two) and ALL (right two) classes.

Similarly, Figure 7c highlights the distinct behavior between the Malaria and Normal classes, particularly in their *β*_0_(50) values. In the binary images *𝒳*_50_, the Malaria class typically exhibits only a few connected components, whereas the Normal class develops 20–30 components. This clear contrast is evident in the sample images shown in Figure 9, clearly illustrating the difference in connectivity between the two classes.

**Fig 9:**
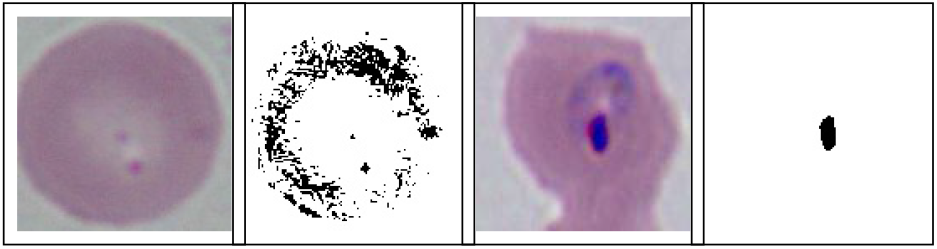
Sample single cell images and their binary *𝒳*_50_ images for Normal (left two) and Malaria (right two) classes.

## 5 Conclusion

In this work, we introduced topological machine learning methods to the domain of cytomorphology and demonstrated their potential in diagnosing critical blood diseases and disorders, including ALL, AML, malaria, and babesiosis. By incorporating topological features into deep learning frameworks, we achieved significant improvements in diagnostic accuracy and model robustness, particularly in data-limited settings where traditional deep learning models often falter. These results highlight the complementary nature of topological methods and deep learning, offering a pathway to more interpretable and effective diagnostic tools. Future work will focus on expanding the methodology to other hematological and rare diseases, exploring real-time integration into clinical workflows, and leveraging advancements in self-supervised learning to further mitigate the challenges posed by limited labeled datasets.

## Data Availability

All data produced are available online at
https://www.cancerimagingarchive.net/collection/aml-cytomorphology_mll_helmholtz/
https://www.kaggle.com/datasets/sizlingdhairya1/all-idb-images

## Acknowledgments

This work was partially supported by the National Science Foundation under grants DMS-2202584, 2229417, and DMS-2220613 and by Simons Foundation under grant # 579977. The authors acknowledge the <u>Texas Advanced Computing Center</u> (TACC) at UT Austin for computational resources which contributed to the research results reported within this paper.

## Footnotes

3 https://anonymous.4open.science/r/TopoVita-X-555C

